# The Q-IMPROvE (Queensland-IMplementation of PRecision Oncology in brEast cancer) pilot study

**DOI:** 10.1101/2023.02.21.23286199

**Authors:** Amy E McCart Reed, Therese McCurry, Georgina Hollway, Haidar Al-Saig, Vladimir Andelkovic, Katharine Cuff, Margaret Cummings, David Fairbairn, Po-ling Inglis, Gillian Jagger, Helene Johanson, Lauren Kalinowski, Olga Kondrashova, Lambros T. Koufariotis, Anna Kuchel, Rahul Ladwa, Chiyan Lau, Ben Lundie, Helen Mar Fan, Nicole McCarthy, Kathryn Middleton, Kowsalya Murugappan, Mark Nalder, Colleen Niland, Michelle K Nottage, Kenneth J O’Byrne, John V Pearson, Kate Roberts, Gorane Santamaria Hormaechea, Cameron Snell, Karin Steinke, Aneta Suder, Diana Tam, Euan Walpole, Natasha Woodward, Clement Wong, Ho Yi Wong, Wen Xu, Peter T Simpson, Nicola Waddell, Sunil R Lakhani

## Abstract

**Background:** The cancer genomics field has embraced the advent of precision oncology, and vast volumes of data have been mined for biomarkers of drug actionability. While some cancers, such as lung cancer, have detailed panels of actionable genomic biomarkers, sequencing panels have been less useful in breast cancer given its large number of cancer driver genes mutated at a relatively low frequency. Furthermore, mutation signatures have potential to assist in identifying homologous recombination deficient tumours for targeting with PARP inhibitor therapy.

**Patients and Methods:** To investigate whether whole genome sequencing could benefit breast cancer patients we initiated the Q-IMPROvE (Queensland-IMplementation of PRecision Oncology in brEast cancer) prospective pilot study. We report the analysis of matched tumour and normal genomes of 28 high-risk breast cancer patients undergoing treatment in the neo-adjuvant setting.

**Results:** Using whole genome sequencing, we detected actionable events that would otherwise not have been identified. A quarter of patients demonstrated a defect in homologous recombination DNA repair using the HRDetect and HRD scores. Germline variants of importance (*BRCA1, CHEK2*) were identified in two patients that did not meet clinical guidelines for germline genetic testing. Somatically, *TP53* and *PIK3CA* were the most commonly mutated breast cancer driver genes.

**Conclusions:** We have demonstrated the benefit of whole genome sequencing of both the tumour and germline for breast cancer patients otherwise not meeting clinical criteria for genetic health referrals.

## Main

Most breast cancer driver genes are mutated in less than 5% of cancers resulting in significant interpatient heterogeneity [1, 2], and thus at the driver gene level, Stephens *et al* consider that most primary breast cancers are distinct [2]. The genomics of breast cancer has been comprehensively studied using whole genome sequencing (WGS) of 560 breast cancers [3]. Saturation analyses predict that based on the number of samples now sequenced, most cancer genes implicated in 2% or more breast cancers will have been identified [4]. Even so, a comprehensive effort by Yates *et al*. [5] acknowledges that many more low-frequency cancer genes remain to be discovered, because there is emerging evidence that metastatic, pre-treated, and special histopathologic subtypes of cancers are genomically distinct from the “general” breast cancer population. Identification of these subtypes allows implementation of specific therapies for the different types of breast cancers, enabling each patient to become an ‘n-of-one’ trial, with their response to therapy informing the treatment of future patients. Increasing numbers of tools are emerging to facilitate the matching of genomic alterations and therapies, including for example, OncoKB [6] and PanDrugs [7], while the MD Anderson program [8] is feeding back ‘sequence-drug’ matching data into the public arena through their Precision Cancer Therapy interface. By end 2015, 39 gene targets with matched FDA-approved therapies were noted in an extensive review of precision oncology [9]; and, as of 2022, OncoKB notes 43 genes at Level 1 with approval, and 11 resistance genes (Level R1/R2), across all cancer types.

In some cancer types, a small panel of pre-selected genes can be sequenced to identify mutation status, and deduce therapeutic strategies (e.g. lung, colorectal cancer). Whole exome sequencing encompassing all of the genome’s coding regions can also be useful, however in cancer types such as breast, it can be less reliable for assessing copy number alterations and will miss structural variations occurring outside of exons [10]. WGS allows us to interrogate gene fusions and larger structural rearrangements [11], and perhaps most importantly for breast cancer, to determine mutation signatures [12]. Certain mutation signatures have potential as diagnostic tools. For example, mutation signatures associated with a defective homologous recombination (HR) DNA repair pathway, which is crucial for maintaining genome stability, can be more robust at predicting HR deficiency (HRD) than the analysis of genetic alterations in HR genes alone (e.g. *BRCA1, BRCA2, RAD51C, RAD51D, PALB2* etc) [13]. HRD enhances sensitivity to DNA-damaging chemotherapeutics, making the associated mutation signature a predictive biomarker. The HRDetect tool [14] was developed in breast cancer and combines several types of mutational signatures derived from WGS to predict HR deficiency (HRD) and thus sensitivity to DNA-damaging chemotherapy (for example anthracyclines or platinum-based therapies). There is increasing evidence of HRDetect score applicability in the clinic; it is tolerant of both low tumour cellularity (as low as 13%) and high mutation burden, and proven in the neoadjuvant setting [14, 15]. Moreover, the OlympiA trial has confirmed that in high-risk early breast cancer (HER2-negative breast cancer and germline *BRCA1* or *BRCA2* pathogenic or likely pathogenic variants) there is a significant improvement in disease free survival with the adjuvant addition of the PARP inhibitor, olaparib [16]. Indeed, the importance of germline variants in seemingly non-familial cancer patients is just coming to light, and expanded germline DNA testing in the Memorial Sloan Kettering breast cancer cohort not meeting clinical criteria for germline testing identified a pathogenic or likely pathogenic germline variant in 17.5% cases [17].

We present a pilot study to examine the potential benefit of the implementation of WGS in the breast cancer care pathway in Queensland, Australia.

## Methods

Ethical approval for this pilot study was granted by the Royal Brisbane and Women’s Hospital (HREC/2019/QRBW/48171) and The University of Queensland (2020000203). The registered study (ACTRN12621001285842) was funded as part of Queensland Genomics, a state-wide program to implement genomics into public healthcare [18]. Following a diagnosis of breast cancer, and a decision to undergo neo-adjuvant chemotherapy (as per local standard of care protocols), patients provided informed consent and were recruited to the study. At the time of surgical clip insertion, three 16-gauge core biopsies were taken and collected into RNALater (ThermoFisher Scientific, Melbourne, Australia). DNA and RNA were extracted from tumour cores using the AllPrep kit (Qiagen, Melbourne, Australia). DNA from baseline bloods collected in EDTA tubes was extracted using either the chemagic360™ instrument (PerkinElmer, ThermoFisher Scientific) and Janus extraction kit (PerkinElmer) or the QIASymphony instrument and dedicated kit (Qiagen).

Samples were sequenced by BGI Australia using PE150 chemistry on a DNBSeq-G400 sequencer to a targeted minimum read depth of 30x coverage for normal and 60x for tumour DNA. Comprehensive variant analysis was performed by genomiQa (Brisbane, Australia). Sequence data was aligned to the human genome (GRCh37/hg19) using BWA-MEM, single nucleotide variants (SNVs) and indels were identified using GATK [19] and qSNP [20]. Copy number alterations (CNAs) and tumour content were called using ASCAT [21]. Variants were classified using the ACMG [22] and AMP/ASCO/CAP [23] guidelines. Reporting of somatic variants was restricted to n=261 genes selected based on a literature review including reported breast and other cancer driver genes [3, 24, 25], and germline variants to 6 genes based on current local standard for testing and thus clinically actionable (**SupTable 1**). Pharmacogenomic loci assessed in the germline sequences for each patient were also noted [26].

**Table 1.** Breast cancer variants of interest reviewed in the Q-IMPROvE study.

| **ENSEMBL Gene ID** | **Gene Symbol** | **Somatic/Germline** |
| --- | --- | --- |
| ENSG00000132142 | ACACA | Somatic |
| ENSG00000184009 | ACTG1 | Somatic |
| ENSG00000077080 | ACTL6B | Somatic |
| ENSG00000138071 | ACTR2 | Somatic |
| ENSG00000115091 | ACTR3 | Somatic |
| ENSG00000049192 | ADAMTS6 | Somatic |
| ENSG00000078295 | ADCY2 | Somatic |
| ENSG00000173020 | ADRBK1 | Somatic |
| ENSG00000155966 | AFF2 | Somatic |
| ENSG00000180772 | AGTR2 | Somatic |
| ENSG00000142208 | AKT1 | Somatic |
| ENSG00000204673 | AKT1S1 | Somatic |
| ENSG00000105221 | AKT2 | Somatic |
| ENSG00000161203 | AP2M1 | Somatic |
| ENSG00000134982 | APC | Somatic |
| ENSG00000184945 | AQP12A | Somatic |
| ENSG00000143761 | ARF1 | Somatic |
| ENSG00000160007 | ARHGAP35 | Somatic |
| ENSG00000141522 | ARHGDIA | Somatic |
| ENSG00000117713 | ARID1A | Somatic |
| ENSG00000049618 | ARID1B | Somatic |
| ENSG00000111229 | ARPC3 | Somatic |
| ENSG00000171456 | ASXL1 | Somatic |
| ENSG00000156802 | ATAD2 | Somatic |
| ENSG00000123268 | ATF1 | Somatic |
| ENSG00000149311 | ATM | Somatic and Germline (only variant ATM c.7271T>G) |
| ENSG00000175054 | ATR | Somatic |
| ENSG00000085224 | ATRX | Somatic |
| ENSG00000103126 | AXIN1 | Somatic |
| ENSG00000163930 | BAP1 | Somatic |
| ENSG00000183337 | BCOR | Somatic |
| ENSG00000157764 | BRAF | Somatic |
| ENSG00000012048 | BRCA1 | Somatic and Germline |
| ENSG00000139618 | BRCA2 | Somatic and Germline |
| ENSG00000156970 | BUB1B | Somatic |
| ENSG00000135932 | CAB39 | Somatic |
| ENSG00000102547 | CAB39L | Somatic |
| ENSG00000179218 | CALR | Somatic |
| ENSG00000152495 | CAMK4 | Somatic |
| ENSG00000142453 | CARM1 | Somatic |
| ENSG00000064012 | CASP8 | Somatic |
| ENSG00000130940 | CASZ1 | Somatic |
| ENSG00000067955 | CBFB | Somatic |
| ENSG00000114423 | CBLB | Somatic |
| ENSG00000110092 | CCND1 | Somatic |
| ENSG00000112576 | CCND3 | Somatic |
| ENSG00000105173 | CCNE1 | Somatic |
| ENSG00000118816 | CCNI | Somatic |
| ENSG00000117877 | CD3EAP | Somatic |
| ENSG00000039068 | CDH1 | Somatic |
| ENSG00000170312 | CDK1 | Somatic |
| ENSG00000123374 | CDK2 | Somatic |
| ENSG00000135446 | CDK4 | Somatic |
| ENSG00000105810 | CDK6 | Somatic |
| ENSG00000124762 | CDKN1A | Somatic |
| ENSG00000111276 | CDKN1B | Somatic |
| ENSG00000147889 | CDKN2A | Somatic |
| ENSG00000147883 | CDKN2B | Somatic |
| ENSG00000172757 | CFL1 | Somatic |
| ENSG00000183765 | CHEK2 | Somatic and Germline<br>(truncating only) |
| ENSG00000079432 | CIC | Somatic |
| ENSG00000141367 | CLTC | Somatic |
| ENSG00000088038 | CNOT3 | Somatic |
| ENSG00000005339 | CREBBP | Somatic |
| ENSG00000204435 | CSNK2B | Somatic |
| ENSG00000102974 | CTCF | Somatic |
| ENSG00000044115 | CTNNA1 | Somatic |
| ENSG00000163131 | CTSS | Somatic |
| ENSG00000257923 | CUX1 | Somatic |
| ENSG00000121966 | CXCR4 | Somatic |
| ENSG00000070190 | DAPP1 | Somatic |
| ENSG00000129187 | DCTD | Somatic |
| ENSG00000175197 | DDIT3 | Somatic |
| ENSG00000119772 | DNMT3A | Somatic |
| ENSG00000188641 | DPYD | Somatic |
| ENSG00000108861 | DUSP3 | Somatic |
| ENSG00000101412 | E2F1 | Somatic |
| ENSG00000130159 | ECSIT | Somatic |
| ENSG00000203734 | ECT2L | Somatic |
| ENSG00000146648 | EGFR | Somatic |
| ENSG00000151247 | EIF4E | Somatic |
| ENSG00000100393 | EP300 | Somatic |
| ENSG00000141736 | ERBB2 | Somatic |
| ENSG00000065361 | ERBB3 | Somatic |
| ENSG00000178568 | ERBB4 | Somatic |
| ENSG00000175595 | ERCC4 | Somatic |
| ENSG00000091831 | ESR1 | Somatic |
| ENSG00000117560 | FASLG | Somatic |
| ENSG00000109670 | FBXW7 | Somatic |
| ENSG00000158815 | FGF17 | Somatic |
| ENSG00000070388 | FGF22 | Somatic |
| ENSG00000111241 | FGF6 | Somatic |
| ENSG00000077782 | FGFR1 | Somatic |
| ENSG00000066468 | FGFR2 | Somatic |
| ENSG00000129514 | FOXA1 | Somatic |
| ENSG00000118689 | FOXO3 | Somatic |
| ENSG00000114861 | FOXP1 | Somatic |
| ENSG00000170324 | FRMPD2 | Somatic |
| ENSG00000128242 | GAL3ST1 | Somatic |
| ENSG00000107485 | GATA3 | Somatic |
| ENSG00000156049 | GNA14 | Somatic |
| ENSG00000087460 | GNAS | Somatic |
| ENSG00000127928 | GNGT1 | Somatic |
| ENSG00000204175 | GPRIN2 | Somatic |
| ENSG00000132522 | GPS2 | Somatic |
| ENSG00000177885 | GRB2 | Somatic |
| ENSG00000120251 | GRIA2 | Somatic |
| ENSG00000155974 | GRIP1 | Somatic |
| ENSG00000082701 | GSK3B | Somatic |
| ENSG00000196126 | HLA-DRB1 | Somatic |
| ENSG00000174775 | HRAS | Somatic |
| ENSG00000197915 | HRNR | Somatic |
| ENSG00000166598 | HSP90B1 | Somatic |
| ENSG00000140443 | IGF1R | Somatic |
| ENSG00000147168 | IL2RG | Somatic |
| ENSG00000113520 | IL4 | Somatic |
| ENSG00000136244 | IL6 | Somatic |
| ENSG00000160712 | IL6R | Somatic |
| ENSG00000134352 | IL6ST | Somatic |
| ENSG00000198001 | IRAK4 | Somatic |
| ENSG00000133124 | IRS4 | Somatic |
| ENSG00000123104 | ITPR2 | Somatic |
| ENSG00000143603 | KCNN3 | Somatic |
| ENSG00000147050 | KDM6A | Somatic |
| ENSG00000055609 | KMT2C | Somatic |
| ENSG00000167548 | KMT2D | Somatic |
| ENSG00000133703 | KRAS | Somatic |
| ENSG00000182866 | LCK | Somatic |
| ENSG00000100097 | LGALS1 | Somatic |
| ENSG00000198799 | LRIG2 | Somatic |
| ENSG00000169032 | MAP2K1 | Somatic |
| ENSG00000034152 | MAP2K3 | Somatic |
| ENSG00000065559 | MAP2K4 | Somatic |
| ENSG00000108984 | MAP2K6 | Somatic |
| ENSG00000095015 | MAP3K1 | Somatic |
| ENSG00000135341 | MAP3K7 | Somatic |
| ENSG00000100030 | MAPK1 | Somatic |
| ENSG00000109339 | MAPK10 | Somatic |
| ENSG00000102882 | MAPK3 | Somatic |
| ENSG00000107643 | MAPK8 | Somatic |
| ENSG00000050748 | MAPK9 | Somatic |
| ENSG00000119487 | MAPKAP1 | Somatic |
| ENSG00000135679 | MDM2 | Somatic |
| ENSG00000112282 | MED23 | Somatic |
| ENSG00000133895 | MEN1 | Somatic |
| ENSG00000079277 | MKNK1 | Somatic |
| ENSG00000099875 | MKNK2 | Somatic |
| ENSG00000076242 | MLH1 | Somatic |
| ENSG00000130396 | MLLT4 | Somatic |
| ENSG00000095002 | MSH2 | Somatic |
| ENSG00000173531 | MST1 | Somatic |
| ENSG00000118513 | MYB | Somatic |
| ENSG00000136997 | MYC | Somatic |
| ENSG00000172936 | MYD88 | Somatic |
| ENSG00000008130 | NADK | Somatic |
| ENSG00000158747 | NBL1 | Somatic |
| ENSG00000158092 | NCK1 | Somatic |
| ENSG00000124151 | NCOA3 | Somatic |
| ENSG00000141027 | NCOR1 | Somatic |
| ENSG00000196712 | NF1 | Somatic |
| ENSG00000186575 | NF2 | Somatic |
| ENSG00000104825 | NFKBIB | Somatic |
| ENSG00000134259 | NGF | Somatic |
| ENSG00000106100 | NOD1 | Somatic |
| ENSG00000148400 | NOTCH1 | Somatic |
| ENSG00000134250 | NOTCH2 | Somatic |
| ENSG00000213281 | NRAS | Somatic |
| ENSG00000166368 | OR2D2 | Somatic |
| ENSG00000179468 | OR9A2 | Somatic |
| ENSG00000130669 | PAK4 | Somatic |
| ENSG00000083093 | PALB2 | Somatic and Germline<br>(truncating only) |
| ENSG00000102699 | PARP4 | Somatic |
| ENSG00000075891 | PAX2 | Somatic |
| ENSG00000163939 | PBRM1 | Somatic |
| ENSG00000134853 | PDGFRA | Somatic |
| ENSG00000152256 | PDK1 | Somatic |
| ENSG00000108518 | PFN1 | Somatic |
| ENSG00000156531 | PHF6 | Somatic |
| ENSG00000078142 | PIK3C3 | Somatic |
| ENSG00000121879 | PIK3CA | Somatic |
| ENSG00000105851 | PIK3CG | Somatic |
| ENSG00000145675 | PIK3R1 | Somatic |
| ENSG00000117461 | PIK3R3 | Somatic |
| ENSG00000115020 | PIKFYVE | Somatic |
| ENSG00000127445 | PIN1 | Somatic |
| ENSG00000164093 | PITX2 | Somatic |
| ENSG00000123739 | PLA2G12A | Somatic |
| ENSG00000182621 | PLCB1 | Somatic |
| ENSG00000124181 | PLCG1 | Somatic |
| ENSG00000122512 | PMS2 | Somatic |
| ENSG00000172531 | PPP1CA | Somatic |
| ENSG00000137713 | PPP2R1B | Somatic |
| ENSG00000057657 | PRDM1 | Somatic |
| ENSG00000046889 | PREX2 | Somatic |
| ENSG00000162409 | PRKAA2 | Somatic |
| ENSG00000181929 | PRKAG1 | Somatic |
| ENSG00000114302 | PRKAR2A | Somatic |
| ENSG00000166501 | PRKCB | Somatic |
| ENSG00000171862 | PTEN | Somatic |
| ENSG00000179295 | PTPN11 | Somatic |
| ENSG00000134242 | PTPN22 | Somatic |
| ENSG00000153707 | PTPRD | Somatic |
| ENSG00000188060 | RAB42 | Somatic |
| ENSG00000136238 | RAC1 | Somatic |
| ENSG00000132155 | RAF1 | Somatic |
| ENSG00000144118 | RALB | Somatic |
| ENSG00000139687 | RB1 | Somatic |
| ENSG00000147274 | RBMX | Somatic |
| ENSG00000132677 | RHBG | Somatic |
| ENSG00000067560 | RHOA | Somatic |
| ENSG00000137275 | RIPK1 | Somatic |
| ENSG00000143622 | RIT1 | Somatic |
| ENSG00000156313 | RPGR | Somatic |
| ENSG00000117676 | RPS6KA1 | Somatic |
| ENSG00000177189 | RPS6KA3 | Somatic |
| ENSG00000141564 | RPTOR | Somatic |
| ENSG00000159216 | RUNX1 | Somatic |
| ENSG00000181555 | SETD2 | Somatic |
| ENSG00000115524 | SF3B1 | Somatic |
| ENSG00000175793 | SFN | Somatic |
| ENSG00000160691 | SHC1 | Somatic |
| ENSG00000155926 | SLA | Somatic |
| ENSG00000117394 | SLC2A1 | Somatic |
| ENSG00000175387 | SMAD2 | Somatic |
| ENSG00000141646 | SMAD4 | Somatic |
| ENSG00000127616 | SMARCA4 | Somatic |
| ENSG00000115904 | SOS1 | Somatic |
| ENSG00000065526 | SPEN | Somatic |
| ENSG00000161011 | SQSTM1 | Somatic |
| ENSG00000101972 | STAG2 | Somatic |
| ENSG00000170581 | STAT2 | Somatic |
| ENSG00000168610 | STAT3 | Somatic |
| ENSG00000118046 | STK11 | Somatic |
| ENSG00000183735 | TBK1 | Somatic |
| ENSG00000177565 | TBL1XR1 | Somatic |
| ENSG00000135111 | TBX3 | Somatic |
| ENSG00000121075 | TBX4 | Somatic |
| ENSG00000089225 | TBX5 | Somatic |
| ENSG00000124678 | TCP11 | Somatic |
| ENSG00000168769 | TET2 | Somatic |
| ENSG00000159445 | THEM4 | Somatic |
| ENSG00000156299 | TIAM1 | Somatic |
| ENSG00000244045 | TMEM199 | Somatic |
| ENSG00000067182 | TNFRSF1A | Somatic |
| ENSG00000141510 | TP53 | Somatic and Germline |
| ENSG00000127191 | TRAF2 | Somatic |
| ENSG00000101255 | TRIB3 | Somatic |
| ENSG00000116747 | TROVE2 | Somatic |
| ENSG00000103197 | TSC2 | Somatic |
| ENSG00000109332 | UBE2D3 | Somatic |
| ENSG00000177889 | UBE2N | Somatic |
| ENSG00000124486 | USP9X | Somatic |
| ENSG00000134215 | VAV3 | Somatic |
| ENSG00000100219 | XBP1 | Somatic |
| ENSG00000166913 | YWHAB | Somatic |
| ENSG00000185650 | ZFP36L1 | Somatic |
| ENSG00000147130 | ZMYM3 | Somatic |
| ENSG00000171940 | ZNF217 | Somatic |
| ENSG00000198538 | ZNF28 | Somatic |
| ENSG00000160094 | ZNF362 | Somatic |
| ENSG00000183779 | ZNF703 | Somatic |

SNV mutational signature assignment was performed using deconstructSigs [27] with version2 COSMIC signatures and a signature contribution cut-off of 10%. Rearrangement signature (RS) assignment was performed using YAPSA [28] with previously reported RS signatures [3] and signature contribution cut-off of 10%. Homologous recombination deficiency (HRD) was estimated using scarHRD [29] using allele-specific copy number information determined by ascatNGS, and by HRDetect [14] using HRD sum scores, insertions and deletions with microhomology, SNV and RS signatures. An HRD sum score of ≥ 42 and HRdetect score of >0.7 were used to categorise cancers as HRD. A gene is considered amplified if there are 6 copies or more, or in cases where the overall ploidy is 3.5x or greater, a gene must be amplified 2.5x above the ploidy.

Genomic data was presented as a variant report at regular Molecular Tumour Board (MTB) meetings. We aimed to have representatives from each of the following disciplines present; pathology, molecular oncology, clinical genetics, surgery, imaging, data analysts, cancer genomics specialists, with the requirement that the patient’s medical oncologist (or their representative) be present.

## Results

We present a pilot study and infrastructure pipeline for the clinical implementation of whole genome sequencing for breast cancer patients at three hospitals in Brisbane, Australia. High-risk breast cancer patients due to undergo neo-adjuvant chemotherapy for their breast cancer were recruited to the study and had a blood sample and research core biopsy taken at the time of surgical clip insertion (prior to the commencement of their chemotherapy). These samples underwent WGS, data were analysed and a comprehensive variant report prepared (germline and somatic variants, mutation signatures, HRD scores, pharmacogenomics) and discussed at a multidisciplinary molecular tumour board (MTB) meeting (**Fig1**). Of 29 patients recruited, 28 patients produced sufficient tumour and germline DNA for sequencing, and 26 tumour-germline genome pairs passed quality assurance and proceeded to a variant report. The variant report discussion centred around the identification of potential germline variants that impacted tumour predisposition and known adverse effects to certain treatments (pharmacogenomics), as well as somatic variants that would be considered targetable in the case that the patient required a second line therapy (lack of response to therapy, or a disease recurrence in future). This was discussed in the context of breast cancer biomarkers Estrogen Receptor (ER) and Human Epidermal Growth Factor Receptor (HER2), as indicated from the core biopsy pathology information, and were classed as ER positive (ERpos) or negative (ERneg) or with or without amplification of HER2 (HER2pos, HER2 neg, respectively).

**Figure 1.**
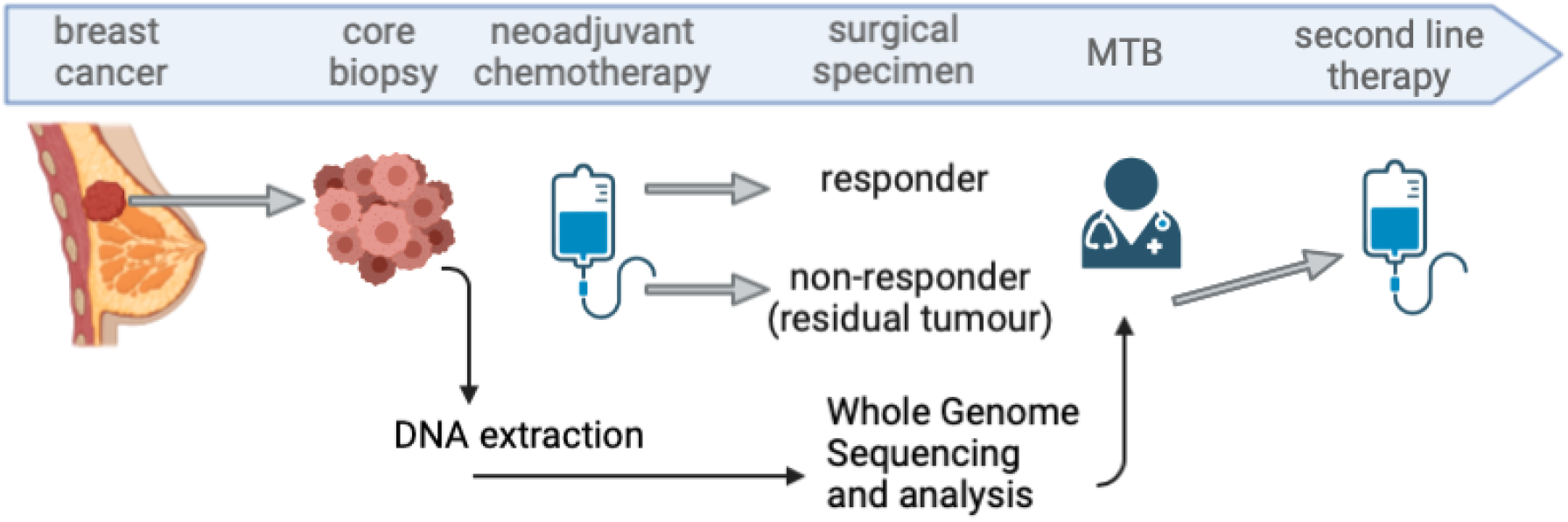
The Q-IMPROvE framework. Following a diagnosis of breast cancer and a decision to proceed with neo-adjuvant therapy, an additional sample (core biopsy) is taken at the time of surgical clip insertion. While the patient undergoes chemotherapy, the pre-therapy tissue and blood sample are sent for DNA extraction and whole genome sequencing. The data is then analysed and returned for discussion at a Molecular Tumour Board (MTB) meeting. Created with BioRender.com.

There were 10 ERpos/HER2neg, five ERpos/HER2pos, seven ERneg/HER2pos and seven TNBC (triple negative breast cancer; ERneg/Progesterone Receptor neg/HER2neg) and the patients ranged in age from 26-69 (median 49), thus reflecting the local clinical guidelines for neo-adjuvant therapy [30], (**Fig2A**). As shown in **Fig2B**, 14/29 cases had a pathologic complete response (pCR) and 15/29 had residual cancer. All ERpos/HER2neg cases had residual cancer (**Fig2C**) and there was a significant enrichment for HER2pos cases in those with pCR (P=0.0008; chi square).

**Figure 2.**
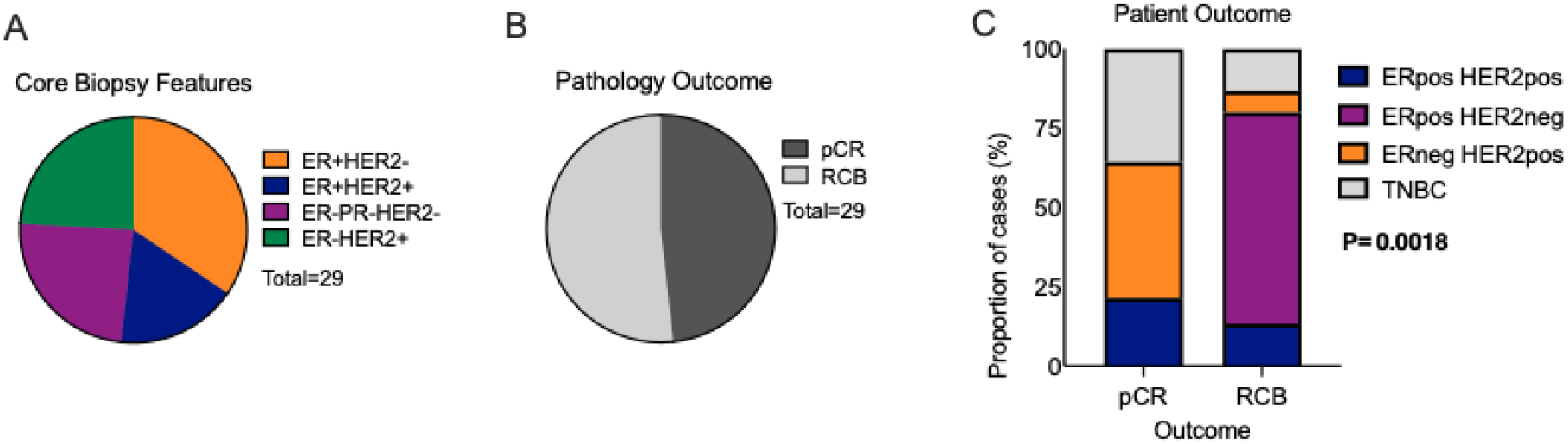
Breast cancer subtypes and outcomes. (A) Of the 29 Q-IMPROvE patients, 11 were HER2pos, there were 15 ERpos and 7 TNBC. (B) 15/29 patients did not achieve a pathologic complete response and had residual cancer burden. (C) The ER pos /HER2neg patients did not respond well to neo-adjuvant therapy and were significantly associated with residual cancer (P=0.0008, chi square). pCR, pathologic complete response; TNBC, triple negative breast cancer.

Of the 29 patients recruited to the study in the funding-mandated timeline, 28 samples were successfully processed into tumour DNA/RNA and germline DNA. The sequencing metrics are detailed in **SupTable 2**. Briefly, the tumour samples were sequenced to an average depth of 73 (55.8 to 90.3); and the normal to 36.5 (33.6 to 40.6). Following sequencing and analysis, 26 samples proceeded with analysis and were prepared into a variant report for discussion, with two samples excluded due to low tumour content.

**Table 2.** Pharmacogenomic variants of interest.

| Gene | Drug | Response | RS number |
| --- | --- | --- | --- |
| *BRINP1* | Trastuzumab | cardiotoxicity<br>(decline in LVEF) | rs10117876 |
| *C10orf11* | Tamoxifen | Recurrence-free<br>survival | rs10509373 |
| *DPYD* | Fluoropyrimidines | Severe toxicity | rs55886062 |
| *EPHA5* | Paclitaxel | Sensory<br>neuropathy | rs7349683 |
| *FGD4* | Paclitaxel | Sensory<br>neuropathy | rs10771973 |
| intergenic_chr14 (*TCL1A*) | Anastrozole,<br>exemestane | Musculoskeletal<br>adverse events | rs11849538 |
| Intergenic_region_of_chr6p22.3 | Trastuzumab | cardiotoxicity<br>(decline in LVEF) | rs4305714 |
| *LDB2* | Trastuzumab | Cardiotoxicity<br>(decline in LVEF) | rs55756123 |
| *RAB22A* | Trastuzumab | cardiotoxicity<br>(decline in LVEF) | rs70755 |
| *SPRR1A/ CACNB4* | Combinations of<br>chemotherapy | Alopecia | rs3820706 |
| *SV2C* | Bevacizumab | Hypertension | rs6453204 |
| *TPD52* | Lapatinib<br>hepatotoxicity | Lapatinib<br>hepatotoxicity | rs7828135 |
| *TRPC6* | Trastuzumab | cardiotoxicity<br>(decline in LVEF) | rs77679196 |
| ZNF613 | Endocrine therapy | Survival | rs8113308 |
| intergenicVariant_chr15 | Anthracycline | Congestive heart failure | rs28714259 |
| regulatoryRegionVariant_chr8/<br>Intergenic region of chr8q21.11 | Anastrozole,<br>exemestane | Breast cancer-free interval | rs13260300 |

### The importance of germline sequencing

An important feature of our study was the germline sequencing, which is not always performed in diagnostic tumour genomic testing. To adhere to our strict ethical oversight, we filtered germline alterations to report only those considered clinically actionable (**SupTable 1**). Eight patients had an existing referral for clinical genetics testing and counselling (Genetic Health Queensland; local clinical guidelines for referral applied and included the Manchester score [31]), two of these patients were known to be *BRCA1* mutation carriers, the other six patients had no known pathogenic alteration (**SupTable 3**). Germline WGS detected the two known *BRCA1* mutations, in addition, two patients were referred to Genetic Health Queensland after Q-IMPROvE WGS analysis identified pathogenic germline *BRCA1* and a *CHEK2* mutation (with copy neutral loss of heterozygosity in the tumour), demonstrating the value of germline sequencing with restricted analysis. An additional patient was also referred to Genetic Health Queensland for accredited germline genetic testing due to a young age of diagnosis and the detection of a somatic *TP53* (C135Y) variant and a copy neutral LOH; the presence of a germline variation would impact the decision to go forward with radiation therapy in the short term, as well as family cancer implications.

**Table 3.**
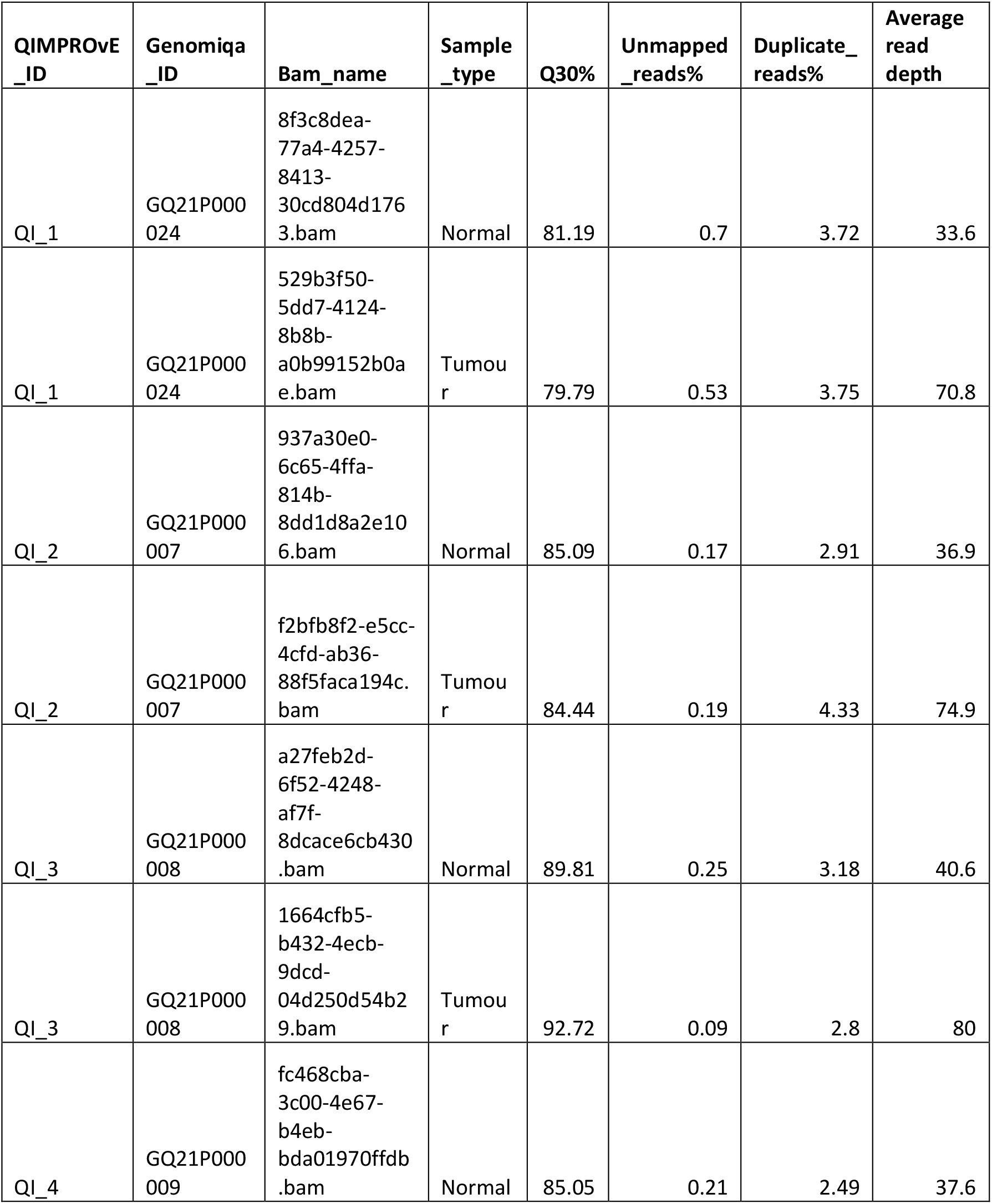
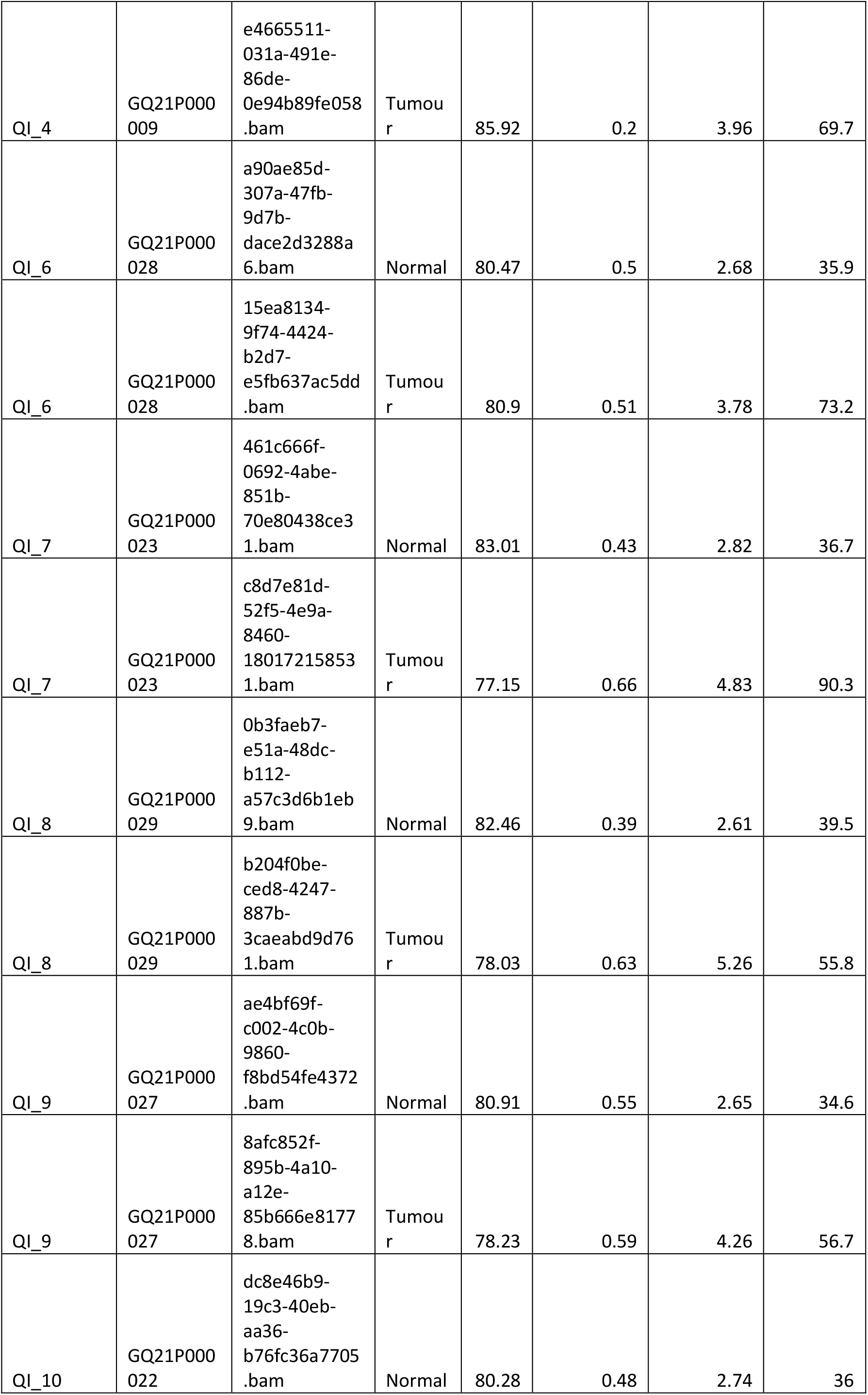
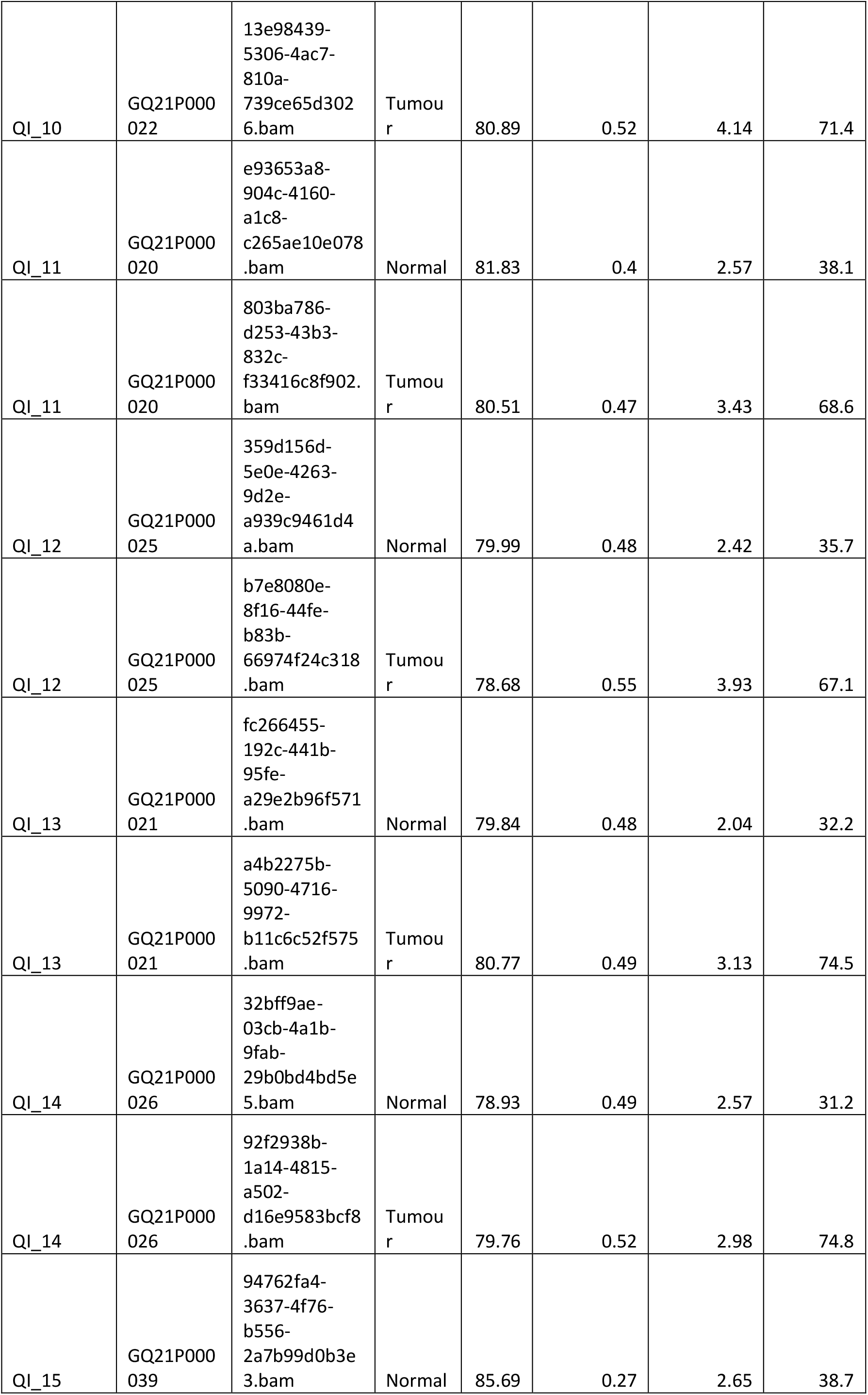
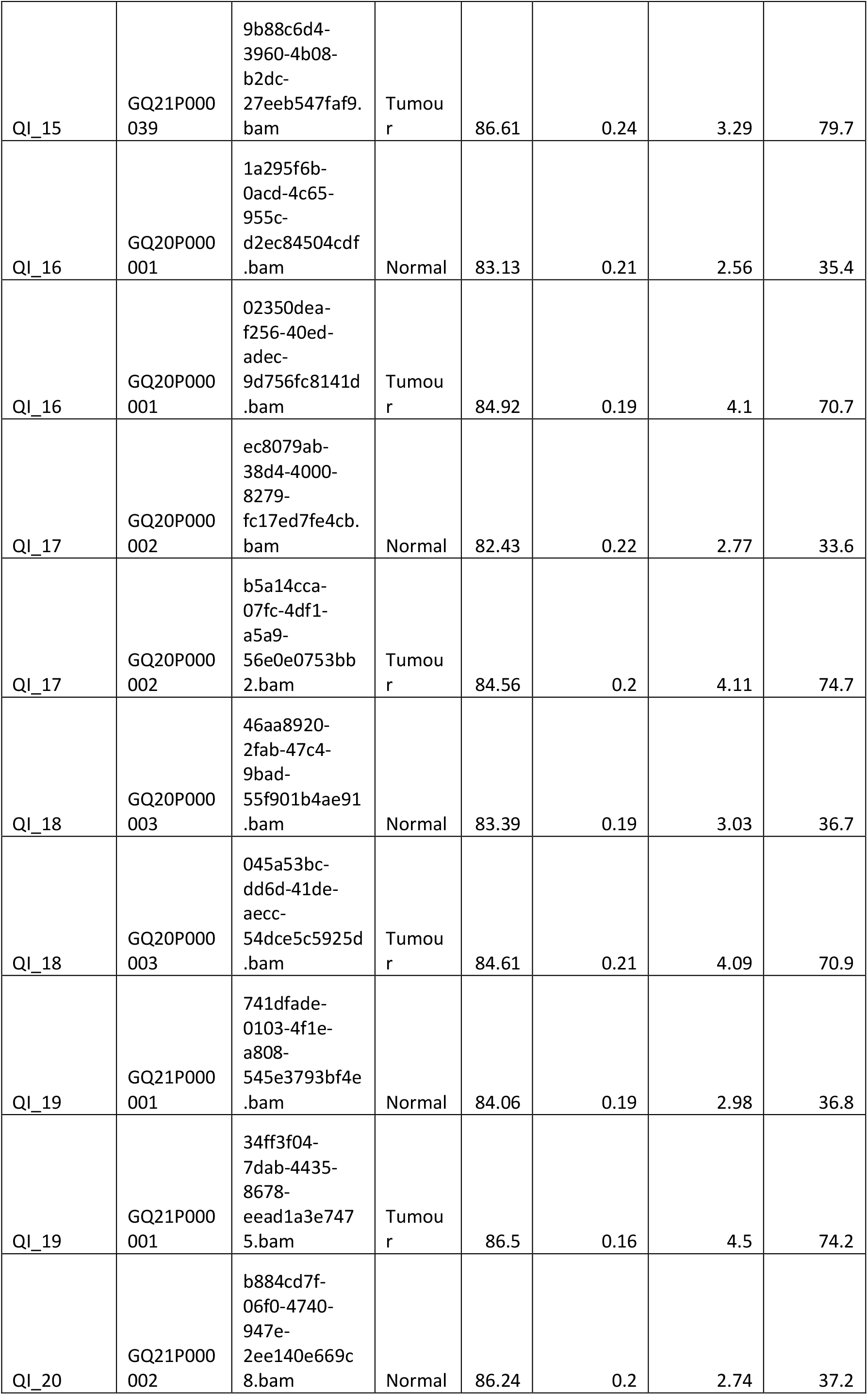
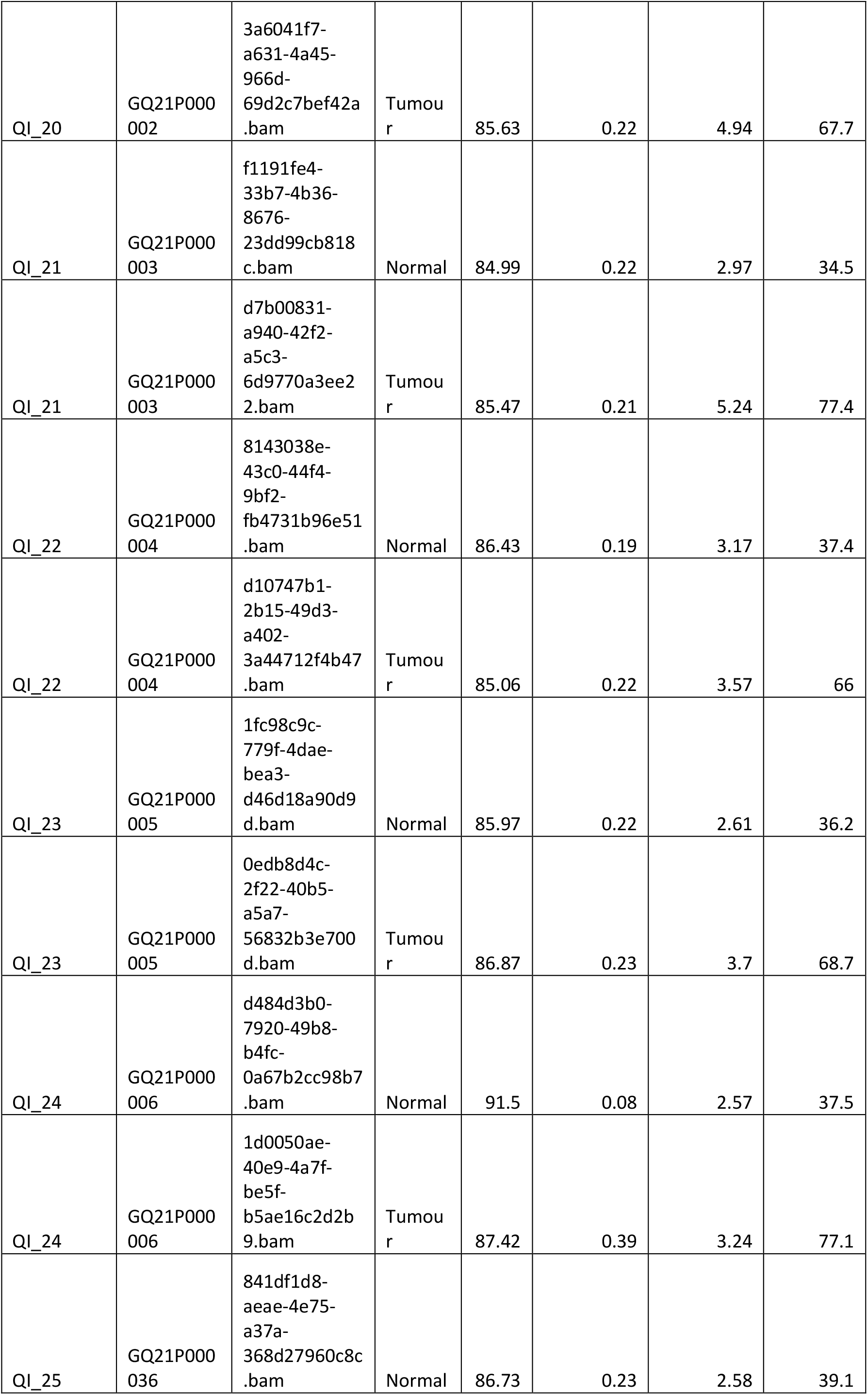
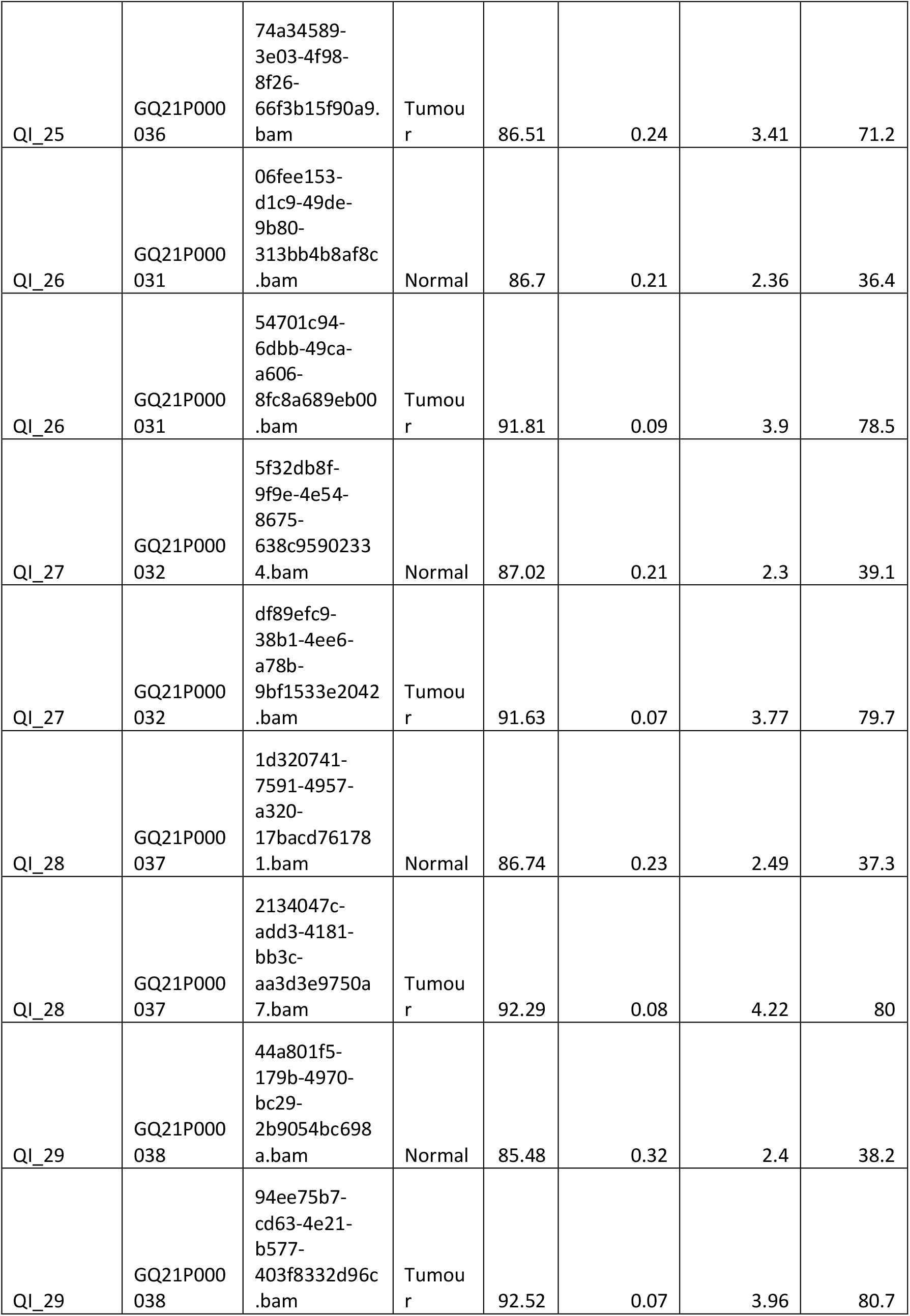
Sequencing Quality Control Data.

Pharmacogenomics is an emerging field that can facilitate the appropriate targeting of existing drugs to patients, in a manner which will reduce potential toxic side effects identified by specific germline variations. Most commonly, a patient’s predisposition to toxicity from capecitabine/fluorouracil can be predicted by investigating the genotype of the *DPYD* gene.

Typically, 10-40% of patients show a severe toxic response to this class of chemotherapeutic [32], and we identified one patient with a heterozygous variation known to impact drug metabolism and mediate toxicity (rs55886062; c.1679 T>G). Crucially this patient had an incomplete response to therapy with residual cancer remaining, and the sequence data informed the selection of an alternative chemotherapeutic to capecitabine, in order to reduce potential toxicity during second-line therapy. We also assessed a number of pharmacogenomic loci as detailed in Low *et al* [26]. However, this data did not yield informative outcomes, for example, three clinically HER2 negative patients had genotypes consistent with trastuzumab/lapatinib sensitivity. While 16 patients (57%) were predicted to have variants associated with neuropathies, our clinical team noted anecdotally that neuropathy is a very common chemotherapy induced side effect, and that these variants would be unlikely to change their management of the patient at this stage.

In summary, the germline analysis identified an additional three instances of actionable alterations, and independently confirmed two existing germline mutations.

### Somatic Alterations: what did we look for, what did we find?

A subtractive analysis of tumour compared to normal was performed to characterise the somatic alterations in each patient (**Fig3**). In addition to mutations, the tumour genomes were assessed for mutation burden, ploidy, mutational signatures, chromosomal rearrangements and copy number alterations. As expected, these breast cancer biopsies did not show a high tumour mutation burden, with a range of 0.4 – 5.7 mutations/Mb (median 1.6), thus none met the 10 mutations/Mb required for predicting pembrolizumab eligibility. The range of ploidy status was 1.6-5, with a median of 3.16.

**Figure 3.**
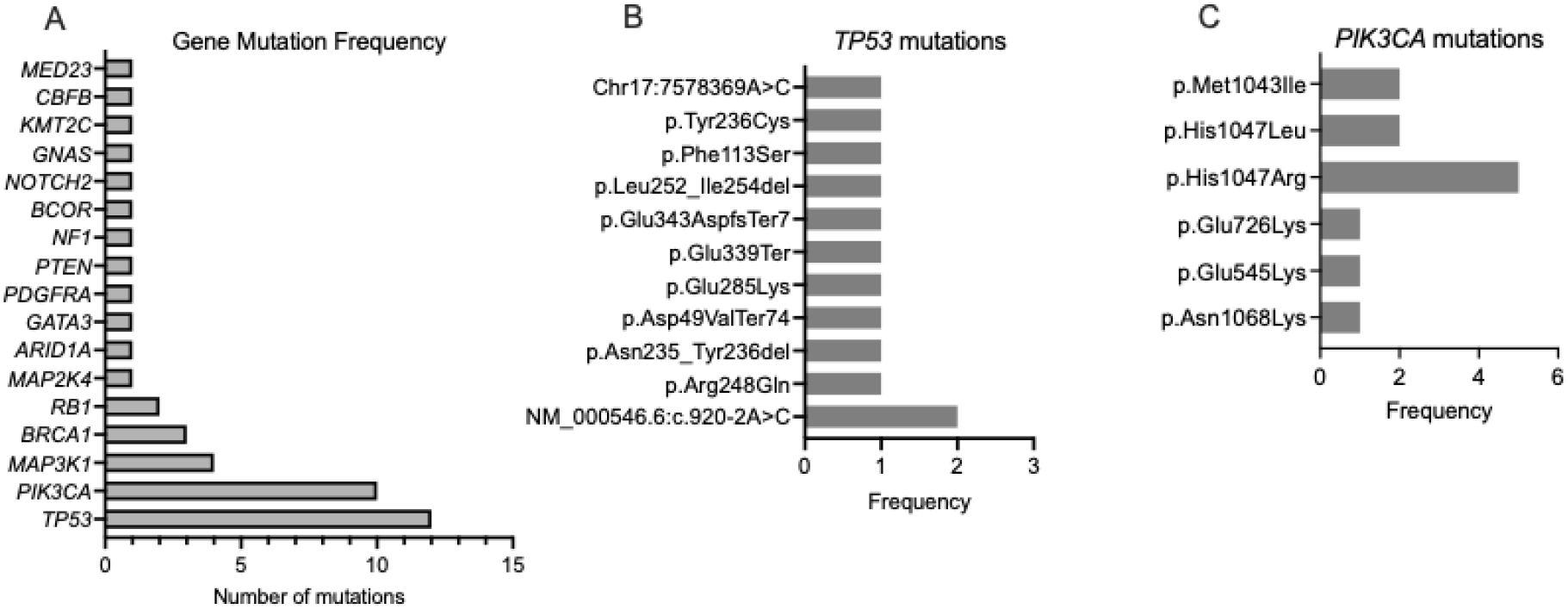
Mutational landscape of Q-IMPROvE cohort. Data per patient, ordered according to standard ER, PR and HER2 status. TMB, tumour mutation burden (mutations/MB). Purity, % tumour cellularity. Ploidy, number of sets of chromosomes. SBS, single base substitution signature, proportion of each substitution signature present per tumour. RS, rearrangement signature, proportion of each rearrangement signature per tumour. CN-LOH, copy neutral– loss of heterozygosity.

We curated a list of 261 breast cancer relevant genes to inform the somatic nucleotide variants and indels included on the report (**SupTable 1**). *TP53* was the most frequently mutated gene (n=13), followed by *PIK3CA* (n=12 across 10 patients), *MAP3K1* (n=4) and *BRCA1* (n=3) (**Fig4A**). *TP53* variations were spread throughout the gene (**Fig4B**) and included three intronic/splice site mutations; these variants were considered as likely oncogenic but of unknown significance in terms of actionability [33]. *PIK3CA* mutations were predictably clustered, with seven at residue 1047 (**Fig4C**); these were classed as a Tier IA mutation classification in ERpos patients.

**Figure 4.**
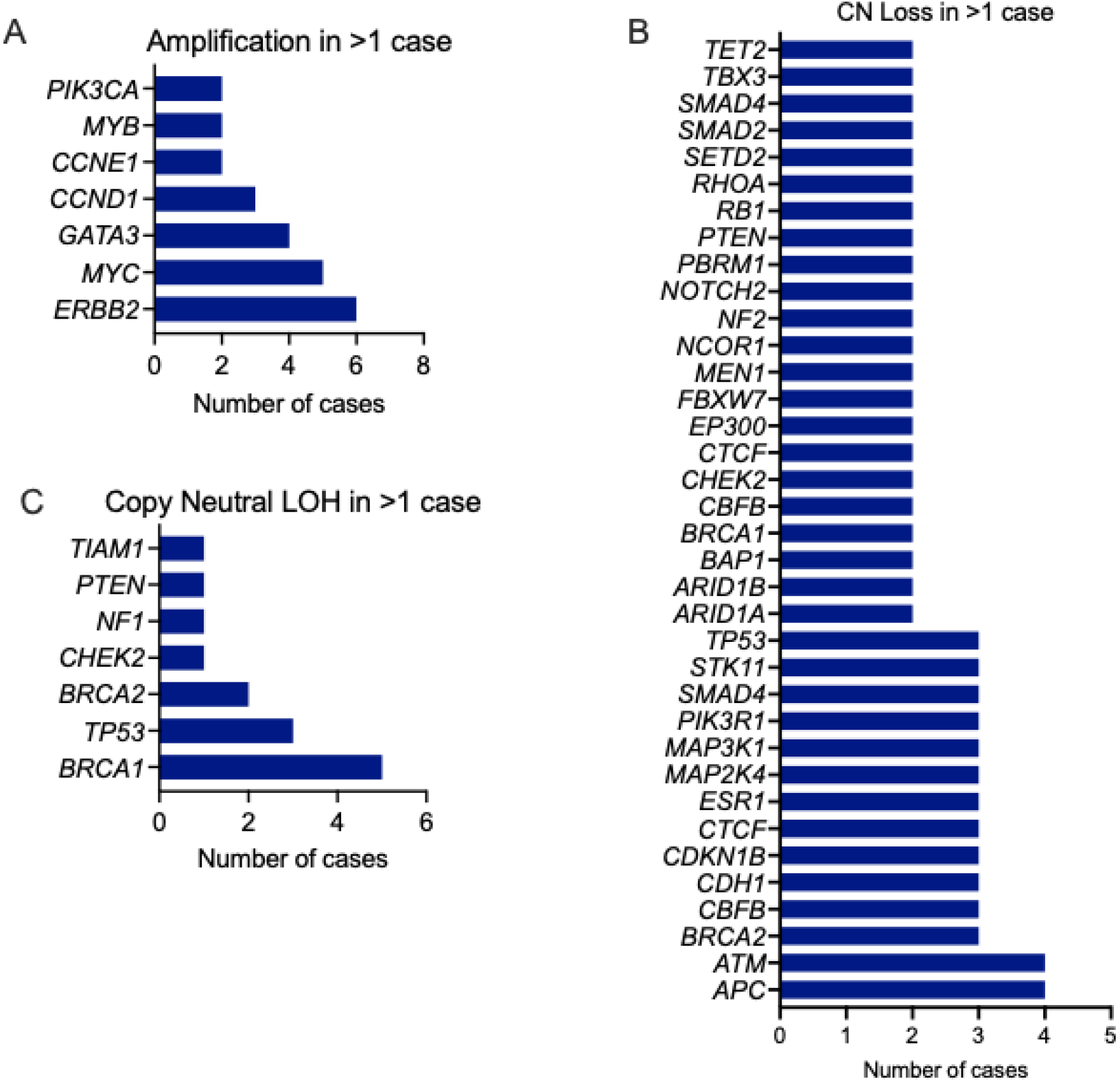
Somatic mutations and Copy number alterations in the Q-IMPROvE cohort. (A) the most frequently mutated genes are plotted, showing the long tail of genes mutated at low frequency. (B) *TP53* mutations were spread across the gene, as expected, while (C) the *PIK3CA* hotspot at amino acid 1047 was highly represented. Recurrent (D) single copy losses, (E) copy neutral Loss of Heterozygosity (CN LOH) and (F) amplifications of well-characterised oncogenes and tumour suppressor genes. (G) Correlation of *ERBB2* copy number from genome sequencing with HER2 Immunohistochemistry from pathology report; amplification is considered to be >6 copies. IHC, immunohistochemistry.

With respect to copy number alterations, **Fig4D** shows the large spread of tumour suppressor genes (n=36) with a single copy loss in two or more patients. An additional 35 genes (**SupTable 4**) demonstrated single copy loss in individual patients only. Copy neutral loss of heterozygosity, as a second hit, was most frequently found in *BRCA1* (n=5 patients) and *TP53* (n=3) as shown in **Fig4E** (**SupTable 4**). *CDKN2A* (p16^INK4a^) and *CDKN2B* (p15^INK4b^), which co-localise on 9p21.3, were found to be homozygously deleted in a single patient, and it is likely that the loss of these well-characterised tumour suppressors impacted tumour development. Frequently amplified genes included *MYC* (n=5 patients), *GATA3* (n=5), the actionable biomarker *CCND1* (n=3), and *CCNE1* (n=2), *MYB* (n=2) and *PIK3CA* (n=2) (**Fig4F; Table 2**) with a high gain reported in an additional 11 genes in individual cases only. *ERBB2* (HER2) was the most recurrently amplified (n=10), the *ERBB2* amplifications were consistent with those cases clinically reported as HER2 amplified by *in situ* hybridisation (**Fig4G**), with the highest *ERBB2* copy number reported to be 83 copies.

**Table 4.** Breast cancer relevant germline findings in the Q-IMPROvE cohort.

| Age<sup>^</sup> | Existing Genetic Health referral for BrCa panel* | Germline Status at recruitment | WGS germline finding | Putative somatic second hit | WGS Somatic finding of interest | New referral to Genetic Health |
| --- | --- | --- | --- | --- | --- | --- |
| 25-35 | Y | None | None | Copy neutral LOH *TP53* | *TP53* p.Cys135Tyr | Yes |
| 25-35 | Y | *BRCA1* mutant | *BRCA1* p.Asn1355Lysfs Ter10 |  | . | . |
| 25-35 | Y | None | None |  | . | . |
| 45-55 | Y | None | None |  | . | . |
| 45-55 | Y | None | None |  | . | . |
| 45-55 | Y | *BRCA1* mutant | *BRCA1* p.Gly1348Asnfs Ter7 | Copy loss *BRCA2* | . | . |
| 45-55 | Y | None | None |  | . | . |
| 45-55 | N | None | *BRCA1* p.Val627SerfsTer4 |  | . | Yes |
| 45-55 | N | None | *CHEK2* p.Gln20Ter | Copy neutral LOH *CHEK2* | . | Yes |
| 60+ | Y | None | None | Copy neutral LOH *BRCA1* | . | . |

<sup>^</sup>patient fits within age bracket indicated;
\*local clinical, germline breast cancer panel comprises: ATM (only variant c.7271T>G); BRCA1; BRCA2; CHEK2 (truncating only); PALB2 (truncating only); TP53.
BrCa, breast cancer; LOH, Loss of heterozygosity; WGS, Whole genome sequencing.

**Table 5.** Frequency of copy number alterations in the Q-IMPROvE cohort.

| Gene | # High Gain | Gene | # Copy Loss | Gene* | # CN LOH | Gene | # HOMD |
| --- | --- | --- | --- | --- | --- | --- | --- |
| *ERBB2* | 6 | *APC* | 4 | *BRCA1* | 5 | *CDKN2A* | 1 |
| *MYC* | 5 | *ATM* | 4 | *TP53* | 3 | *CDKN2B* | 1 |
| *GATA3* | 4 | *BRCA2* | 3 | *BRCA2* | 2 |  |  |
| *CCND1* | 3 | *CBFB* | 3 | *CHEK2* | 1 |  |  |
| *CCNE1* | 2 | *CDH1* | 3 | *NF1* | 1 |  |  |
| *MYB* | 2 | *CDKN1B* | 3 | *PTEN* | 1 |  |  |
| *PIK3CA* | 2 | *CTCF* | 3 | *TIAM1* | 1 |  |  |
| *AKT1* | 1 | *ESR1* | 3 |  |  |  |  |
| ATAD2 | 1 | MAP2K4 | 3 |  |  |  |  |
| BRAF | 1 | MAP3K1 | 3 |  |  |  |  |
| FOXO3 | 1 | PIK3R1 | 3 |  |  |  |  |
| GNAS | 1 | SMAD4 | 3 |  |  |  |  |
| PREX2 | 1 | STK11 | 3 |  |  |  |  |
| RAF1 | 1 | TP53 | 3 |  |  |  |  |
| SLA | 1 | ARID1A | 2 |  |  |  |  |
| TBL1XR1 | 1 | ARID1B | 2 |  |  |  |  |
| TBX4 | 1 | BAP1 | 2 |  |  |  |  |
| TP53 | 1 | BRCA1 | 2 |  |  |  |  |
|  |  | CBFB | 2 |  |  |  |  |
|  |  | CHEK2 | 2 |  |  |  |  |
|  |  | CTCF | 2 |  |  |  |  |
|  |  | EP300 | 2 |  |  |  |  |
|  |  | FBXW7 | 2 |  |  |  |  |
|  |  | MEN1 | 2 |  |  |  |  |
|  |  | NCOR1 | 2 |  |  |  |  |
|  |  | NF2 | 2 |  |  |  |  |
|  |  | NOTCH2 | 2 |  |  |  |  |
|  |  | PBRM1 | 2 |  |  |  |  |
|  |  | PTEN | 2 |  |  |  |  |
|  |  | RB1 | 2 |  |  |  |  |
|  |  | RHOA | 2 |  |  |  |  |
|  |  | SETD2 | 2 |  |  |  |  |
|  |  | SMAD2 | 2 |  |  |  |  |
|  |  | SMAD4 | 2 |  |  |  |  |
|  |  | TBX3 | 2 |  |  |  |  |
|  |  | TET2 | 2 |  |  |  |  |
|  |  | ATRX | 1 |  |  |  |  |
|  |  | AXIN1 | 1 |  |  |  |  |
|  |  | **Gene** | **# Copy Loss** |  |  |  |  |
|  |  | BAP1 | 1 |  |  |  |  |
|  |  | BCOR | 1 |  |  |  |  |
|  |  | BRCA1 | 1 |  |  |  |  |
|  |  | CASP8 | 1 |  |  |  |  |
|  |  | CDH1 | 1 |  |  |  |  |
|  |  | CDKN2A | 1 |  |  |  |  |
|  |  | CHEK2 | 1 |  |  |  |  |
|  |  | CREBBP | 1 |  |  |  |  |
|  |  | EP300 | 1 |  |  |  |  |
|  |  | *ERBB4* | 1 |  |  |  |  |
|  |  | *ERCC4* | 1 |  |  |  |  |
|  |  | *FOXO3* | 1 |  |  |  |  |
|  |  | *KDM6A* | 1 |  |  |  |  |
|  |  | *KMT2C* | 1 |  |  |  |  |
|  |  | *KMT2D* | 1 |  |  |  |  |
|  |  | *MAP2K4* | 1 |  |  |  |  |
|  |  | *MAP3K1* | 1 |  |  |  |  |
|  |  | *MLH1* | 1 |  |  |  |  |
|  |  | *NCOR1* | 1 |  |  |  |  |
|  |  | *NF1* | 1 |  |  |  |  |
|  |  | *NF2* | 1 |  |  |  |  |
|  |  | *PALB2* | 1 |  |  |  |  |
|  |  | *PBRM1* | 1 |  |  |  |  |
|  |  | *PHF6* | 1 |  |  |  |  |
|  |  | *PMS* | 1 |  |  |  |  |
|  |  | *PRDM1* | 1 |  |  |  |  |
|  |  | *PTPRD* | 1 |  |  |  |  |
|  |  | *RHOA* | 1 |  |  |  |  |
|  |  | *SETD2* | 1 |  |  |  |  |
|  |  | *SMAD2* | 1 |  |  |  |  |
|  |  | *SMARCA4* | 1 |  |  |  |  |
|  |  | *STAG2* | 1 |  |  |  |  |
|  |  | *TSC2* | 1 |  |  |  |  |
| * CN LOH reported only for those genes with a mutation also recorded |  |  |  |  |  |  |  |
| CN, copy neutral; HOMD, homozygous deletion; LOH, Loss of heterozygosity. |  |  |  |  |  |  |  |

### Homologous Recombination Deficiency analysis

Mutation signature analysis was used to estimate HRD using HRD Score [34] and HRDetect [14], which are two clinically implementable algorithms used to measure HRD as a proxy for *BRCA1, BRCA2* or other key HR gene dysfunction. These algorithms are incredibly useful, as there are many modes of inactivation of *BRCA1/2*, including mutation, large-scale rearrangements, copy number alterations and promoter methylation; not all variations are known to be pathogenic and no single test can capture all of these potential (epi-)genotypes. For Q-IMPROvE, we required that the criteria for both HRD Score and HRDetect be met to class a patient as HRD. In total, seven cases were determined to be HRD (**Fig5**). Four of these cases had a germline line alteration in *BRCA1/CHEK2*, there was a single somatic *BRCA1* mutation (second/first hit unknown), and two cases of unknown aetiology. In the cohort, there were four cases meeting the HRD Score cut-off (>42) but not the HR Detect (>0.7) and so were classed as HR proficient, but no cases with the converse (**Fig5B**).

### Somatic sequencing informed actionability

Taken together, we found 18 actionable events across 18 patients (69%) through tumour sequencing of 26 patients. Considering currently actionable changes, **Fig5C** summarises the association between those genomic biomarkers and the patient outcome overall; indicating that actionable events may prove important in the instances of tumour recurrence. Notably, an incomplete response to neo-adjuvant therapy in tumours where *PIK3CA* were identified (**Fig5D**) indicating that these patients could respond to alpelisib as a second line therapy to target the *PIK3CA* mutations should they relapse. Another actionable change identified was *CCND1* amplification (n=3), where patients could be triaged to Palbociclib and Avelumab, through a local clinical trial the MoST 10 substudies 23-24 (ACTRN12620000568910). Four patients with residual cancer showed defective homologous recombination DNA repair, indicating that they may be sensitive to olaparib.

**Figure 5.**
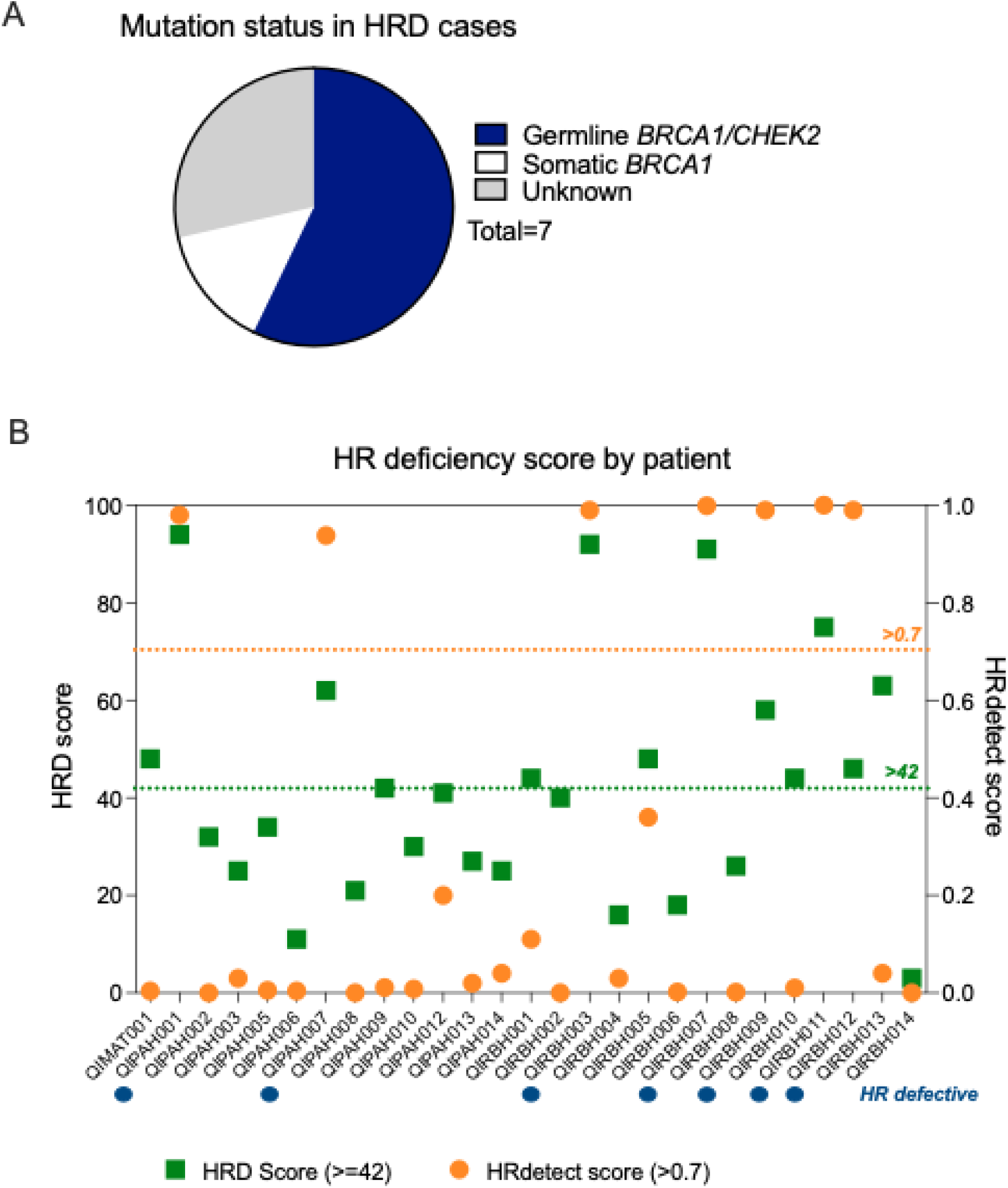
Homologous Recombination Deficiency and other actionable findings across the Q-IMPROvE cohort. (A) the seven HRD cases shown by likely cause. (B) All patients as a function of HRD Sum score (left Y axis) and HRDetect (right Y axis). The blue line indicates, the HRdetect cut-off score (0.7), while the grey line shows the HRD score cut-off (42). Patients marked with a purple circle meet requirements for both tests and are HR defective. (C) summary of actionable changes across the cohort. (D) the relationship between actionable changes and the outcome of neo-adjuvant therapy. No significant enrichment was detected (P=0.1295). pCR, pathologic complete response.

**Figure 6.**
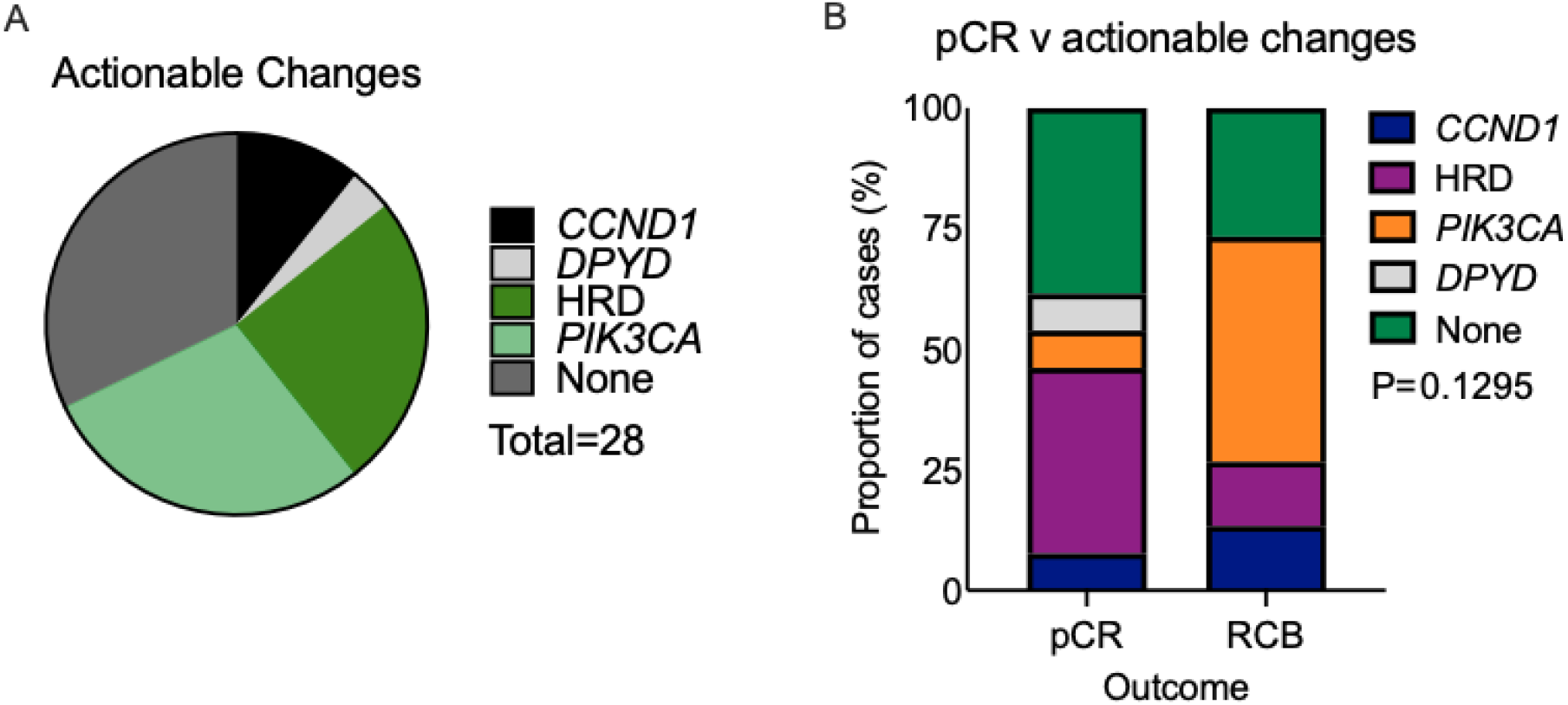
Actionable alterations identified in the Q-IMPROvE study. (A) summary of actionable changes across the cohort. (B) the relationship between actionable changes and the outcome of neo-adjuvant therapy. No significant enrichment was detected (P=0.1295).

## Discussion

We present a clinical framework for the implementation of whole genome sequencing in breast cancer care. We selected tumour-normal paired WGS and accepted its cost limitations in order to prioritise data generation, in particular around germline variants predicting risk and pharmacogenomic toxicities, broad driver gene detection and mutation signature assessment, in particular for HRD assessment. Furthermore, we necessitated the inclusion of paired normal sequencing to eliminate uncertainty around the identification potential germline findings in somatic data. The traditionally long turnaround time of WGS and analysis was offset by our application of the technology in the neo-adjuvant setting, where sequencing and analysis could occur during treatment cycles, which typically take 20-24 weeks [30].

We identified seven cases of HRD in total, with only two of these cases commencing the study with a confirmed pathogenic alteration in a germline breast cancer predisposition gene. Regarding gene specific actionability, we present 12 *PIK3CA* mutations harboured by ten patients; a single case of *DPYD* predicting toxicity; and, three cases of *CCND1* amplification. We confirmed that WGS-derived *ERBB2* copy number status could reliably replace HER2 IHC and *in situ* hybridisation. In 9/28 patients, no actionable changes were identified. While no therapy changes were mandated by our study, it is clear that there is the capacity for WGS to be applied with outcomes benefiting patients around therapy selection, and clinical trial engagement.

That seven of 28 patients (25%) showed HRD, is a strong argument for its routine clinical implementation in breast cancer care. Our application of a virtual panel filter of the germline sequence to restrict findings to those of breast cancer relevance was ethical, useful, and readily implementable. Furthermore, as risk genes, pharmacogenomics and the mutation signature space are all dynamic with novel data likely to be emerging, a whole genome sequence can be reanalysed to include any new loci of interest, representing a future proofing of the data.

We acknowledge the limitations of this study, in its small size and wholly neo-adjuvant setting. Additionally, we accept that the genome of an early breast cancer, pre-treatment, is unlikely to be identical to the genome of a post-treatment tumour being discussed for second-line therapy. However, we considered that the decision to trial WGS in neoadjuvant patients outweighed the potential problems presented by therapy-induced genome alterations.

This study provides evidence to support the introduction of genomics as standard of care in breast cancer management in Australia. Further study is required to determine whether a custom panel or similar could recapitulate this level of data generation and improved cost and turnaround time, and whether the adjuvant treatment setting would also be appropriate.

## Data Availability

All data produced in the present study are available upon reasonable request to the authors

## Acknowledgements

We acknowledge funding from Queensland Genomics (Queensland Health) and the Medical Research Futures Fund, Genomics Health Futures Mission. Thank you to the patients and their families. We thank the many staff across the RBWH, PAH and Mater hospitals that helped facilitate this study. The study was funded by Queensland Health through Queensland Genomics as an Innovation study.

## Author contributions

Study design and infrastructure:, AMR, CS, EW, KOB, NM, NWa, NWo, PTS, SRL.

Patient recruitment co-ordination: GJ, TM

Patient recruitment, clinical oversight, and MTB input: AK, AS, CS, CW, DT, EW, HA, HM, KC, KMi, KMu, KOB, KR, MNa, MNo, NM, NWo, PI, RL, SRL, TM, VA, WX.

Biopsy collection: GSH, KS.

Biopsy review: MC, SRL. Sample processing: HJ.

Pathology oversight and interpretation: BL, CL, CS, DF, LK, MC, SRL.

Variant report design: AMR, GH, HM, LK, NWa.

Data analysis: BL, GH, OK, LK, HYW, JVP, PTS, NWa, AMR.

Critical appraisal, manuscript writing, correction, and approval of final manuscript: all authors.

